# Paediatric traumatic brain injury as a risk factor for psychosis and psychotic symptoms: a systematic review and meta-analysis

**DOI:** 10.1101/2023.02.17.23286118

**Authors:** King-Chi Yau, Grace Revill, Graham Blackman, Madiha Shaikh, Vaughan Bell

## Abstract

**Background:** Psychosis is one of the most disabling psychiatric disorders. Paediatric traumatic brain injury (pTBI) has been cited as a developmental risk factor for psychosis, however this association has never been assessed meta-analytically.

**Methods:** A systematic review and meta-analysis of the association between pTBI and subsequent psychotic disorders/symptoms was performed. The study was pre-registered (CRD42022360772) adopting a random-effects model to estimate meta-analytic odds ratio (OR) and 95% confidence interval (CI) using the Paule–Mandel estimator. Subgroup (study location, study design, psychotic disorder vs subthreshold symptoms, assessment type, and adult vs adolescent onset) and meta-regression (quality of evidence) analyses were also performed. The robustness of findings was assessed through sensitivity analyses. The meta-analysis is available online as a computational notebook with an open dataset.

**Results:** We identified 10 relevant studies and eight were included in the meta-analysis. Based on a pooled sample size of 479,686, the pooled OR for the association between pTBI and psychosis outcomes was 1.80 (95% CI [1.11, 2.95]). There were no subgroup effects and no outliers. Both psychotic disorder and subthreshold symptoms were associated with pTBI. The overall association remained robust after removal of low-quality studies, however the OR reduced to 1.43 (95% CI [1.04, 1.98]). A leave-one-out sensitivity analysis showed the association was robust to removal of all but one study which changed the estimate to marginally non-significant.

**Conclusions:** We report cautious meta-analytic evidence for a positive association between pTBI and future psychosis. New evidence will be key in determining long-term reliability of this finding.

## Introduction

There is consistent evidence indicating that traumatic brain injury (TBI) is associated with an increased risk of adverse neuropsychiatric outcomes in adults, including depression, anxiety, posttraumatic stress symptoms, cognitive impairment, personality change, and neurodegenerative disorders (Carroll et al., 2014; Cnossen et al., 2017; Fleminger, 2008; Perry et al., 2016; Rogers & Read, 2007; Schwartz, Jodis, Breen, & Parker, 2019; van Reekum, Cohen, & Wong, 2000). One association that has proved more controversial, however, has been the link between TBI and psychosis. Although there are clearly cases of post-TBI psychosis (Fujii & Ahmed, 2002), the extent to which TBI is a reliable population risk factor for psychosis has been debated. In a narrative review of the evidence, David and Prince (2005) concluded that it was unlikely brain injury reliably causes psychosis given the published data available at the time. In a subsequent narrative review, Batty et al. (2013) estimated that psychosis following TBI appears to be three times more prevalent than psychotic disorders in the general population. Looking specifically at the association between TBI and schizophrenia in case-control studies, Molloy et al.’s (2011) meta-analysis reported a significant association and, through the inclusion of family studies, suggested this effect was larger in those with a genetic predisposition to psychosis.

Notably, however, the studies considered in these reviews largely examined the impact of adult TBI on later psychosis. Although clearly important, studies that focus solely on adult TBI may miss longer-term associations between TBI that occurs before the age of 18 and an increased risk of psychotic disorders or symptoms later in life. The association between paediatric TBI (pTBI) and psychosis is plausible given what we know about risk factors for psychosis in childhood and adolescence. Key developmental models of psychosis, including the psychosis-proneness-persistence-impairment model (Linscott & van Os, 2013; Os, Linscott, Myin-Germeys, Delespaul, & Krabbendam, 2009) and the developmental risk factor model (Howes & Murray, 2014; Murray, Bhavsar, Tripoli, & Howes, 2017), are based on evidence that adverse experiences that impair typical neurodevelopment can maintain normally transient sub-threshold symptoms of psychosis during adolescence and increase the risk of later transition to psychotic disorders (Rubio, Sanjuan, Florez-Salamanca, & Cuesta, 2012; Trotta, Murray, & Fisher, 2015). It has been suggested that paediatric TBI could be one such neurodevelopmental risk factor (AbdelMalik, Husted, Chow, & Bassett, 2003), but this has never been subjected to systematic review and meta-analysis. Although Molloy et al. (2011) included a subgroup analysis on paediatric TBI cases in their meta-analysis, only three studies were available at the time, indicating a clear need for a more systematic analysis of this issue as new studies have emerged.

Consequently, we conducted a pre-registered systematic review and meta-analysis to determine the association between paediatric TBI and later psychotic disorders or symptoms of psychosis. To the best of our knowledge, this is the first meta-analysis to examine paediatric TBI as a potential risk factor for psychotic disorders or symptoms.

## Methods

The present systematic review and meta-analysis was undertaken and reported in compliance with the Preferred Reporting Items for Systematic Reviews and Meta-Analyses (PRISMA) 2020 guidelines (Page et al., 2021).

### Eligibility criteria

#### Participants

We included studies that recruited participants of any age or gender with a diagnosis of paediatric traumatic brain injury.

#### Exposures

Paediatric traumatic brain injury was defined as an onset of TBI before adulthood (i.e., < 18 years old). Paediatric TBI could be determined by the age of the study population (e.g. children or adolescents with TBIs) or the time of onset of TBI (e.g. adults with a history of paediatric TBI). We included participants with a diagnosis of paediatric TBI based on validated screening tools, structured clinical interviews, medical records reviews, or clinical diagnosis. TBIs with severity ranging from mild (including concussion) to severe were included. For exclusion, we did not select studies where the occurrence of paediatric TBI could not be determined, and when psychotic disorders or symptoms were not measured. In addition, we did not include studies when exposure to TBI could not be differentiated from other non-TBI conditions within a single group.

#### Comparators

Studies with and without comparison groups were included, with no exclusion criteria applied.

#### Outcomes

The main outcome of interest was presence of psychotic disorders or psychotic symptoms based on validated screening tools, psychometric measures, structured clinical interviews, medical records reviews, or clinical diagnosis. Psychotic disorders included schizophrenia and related disorders, whilst psychotic symptoms included psychosis-risk syndromes, psychotic symptoms and psychotic-like experiences. We only included studies where the onset of the psychotic disorder/symptoms occurred after the TBI. We excluded studies reporting only broad neuropsychiatric outcomes (such as behavioural difficulties) without any specific assessment of psychosis.

#### Types of studies included

We included all peer-reviewed primary studies published in English with no date restrictions. The following types of design were included: randomised or non-randomised controlled trials, retrospective or prospective cohort studies, and case-control studies (including nested case-control and family studies). We excluded meta-analyses, systematic reviews, literature reviews, case reports, case series, qualitative studies, opinion pieces, editorials, comments, newsletters, book chapters, and congress papers.

### Information sources and search strategy

The databases of PsycINFO (Ovid) (from 1806 onwards) and MEDLINE (Ovid) (from 1946 onwards) were searched based on the strategy outlined in Table 1 (see Appendix S1 for full search strategy), with the search carried out independently by two reviewers (KCY, GR). Studies were screened according to the above criteria. Prior to the final analysis searches were re-run on 1^st^ December 2022 to identify any further studies that could be included in the review.

**Table 1.** Search strategy.

| Main term | Search term with operator (PsycINFO) | Search term with operator (MEDLINE) |
| --- | --- | --- |
| Traumatic brain injury | (brain injuries/ OR traumatic brain injury/ OR brain concussion/) OR (TBI OR traumatic brain injur* OR brain injur* OR head injur* OR cerebral trauma OR craniocerebral injur* OR concussion* OR skull fracture*).ab,id,ti | (brain injuries/ OR brain injuries, traumatic/ OR brain concussion/) OR (TBI OR traumatic brain injur* OR brain injur* OR head injur* OR cerebral trauma OR craniocerebral injur* OR concussion* OR skull fracture*).ab,kw,ti |
| Psychotic disorders & psychotic symptoms | (psychosis/ OR schizophrenia/) OR (psychosis OR psychotic OR psychotic disorder* OR psychotic exp* OR psychotic?like exp* OR schizophreni* OR delusional disorder* OR delusion* OR hallucinat* OR psychiatric illness* OR psychiatric disorder*).ab,id,ti | (psychotic disorders/ OR schizophrenia/) OR (psychosis OR psychotic OR psychotic disorder* OR psychotic exp* OR psychotic?like exp* OR schizophreni* OR delusional disorder* OR delusion* OR hallucinat* OR psychiatric illness* OR psychiatric disorder*).ab,kw,ti |
| Child | (childhood birth 12 yrs OR preschool age 2 5 yrs OR school age 6 12 yrs OR adolescence 13 17 yrs).ag OR (infan* OR baby* OR babies OR toddler* OR preschool* OR child* OR pediat* OR paediat* OR prepubescen* OR prepuberty* OR puberty OR pubescen* OR teen* OR young* OR youth* OR minors* OR underag* OR juvenile* OR preadolesc* OR adolesc*).ab,id,ti | (infant/ OR child, preschool/ OR child/ OR adolescent/) OR (infan* OR baby* OR babies OR toddler* OR preschool* OR child* OR pediat* OR paediat* OR prepubescen* OR prepuberty* OR puberty OR pubescen* OR teen* OR young* OR youth* OR minors* OR underag* OR juvenile* OR preadolesc* OR adolesc*).ab,kw,ti |

Note. ab = abstract; ag = age group; id = key concepts; kw = keyword heading; ti = title

### Study selection process

Following removal of duplicates, two reviewers (KCY, GR) independently screened the titles and abstracts of all the records retrieved. A third reviewer (VB) was consulted when a consensus could not be reached. Two reviewers (KCY, GR) independently screened the full-text reports based on the above eligibility criteria, and processes of discussion between the two reviewers and consultation with the third reviewer (VB), in the case of disagreement, were held.

### Data extraction process

A data extraction excel sheet was developed by one of the reviewers (KCY). Two reviewers (KCY, GR) independently extracted study characteristics and outcomes from all the included studies, and data were compared. A third reviewer (VB) was consulted when a consensus could not be reached.

### Data items

#### Outcomes

The main outcome was presence of a psychotic disorder or psychotic symptoms including schizophrenia, psychosis, hallucination, paranoia, psychosis-risk syndromes, and psychotic-like experiences. Diagnoses of schizophrenia, psychosis, hallucination, and paranoia based on the *International Statistical Classification of Diseases and Related Health Problems* (ICD), the *Diagnostic and Statistical Manual of Mental Disorders* (DSM), or Feighner et al. (1972) criteria were used. We also extracted sub-threshold symptoms of psychosis including psychosis-risk syndromes (McGlashan, Walsh, & Woods, 2010) and psychotic-like experiences (PLEs) (Lee et al., 2016). For methods of outcome measurement, validated screening tools and psychometric measures (including Prodromal Psychosis Questionnaire – Brief Child Version [PQ-BC] by Karcher et al. (2018)), structured clinical interviews, medical records reviews, and clinical diagnosis were included. Regarding the onset of psychotic disorders or symptoms, anytime time point was eligible (i.e. childhood, adolescence, or adulthood) provided the onset was after paediatric TBI.

Regarding the major outcome data, we primarily extracted the number of participants experiencing psychotic disorders or symptoms after paediatric TBI. When studies used several methods for reporting the relevant data, we followed *a priori* defined rules of decision to select corresponding data. (i) When both the raw number of participants experiencing psychotic disorders or symptoms and the calculated statistics (e.g., incidence rate ratios [IRRs]; odds ratio [ORs]) were available, we extracted the raw number. (ii) When descriptive statistics of interval measures of psychotic disorders or symptoms and the calculated statistics (e.g., *p* values or effect sizes) were available, we extracted the descriptive statistics. (iii) When both non-imputed and imputed data were reported, we chose the imputed. (iv) Lastly, we extracted the set of raw number based on primary analysis of the original study.

Where the required data had not been published (three studies: Lopez et al., 2022; Orlovska et al., 2014; Timonen et al., 2002), authors were contacted for the required information (e.g., asking for total number of participants in the exposure group of paediatric TBI. Two authors responded but only one (Lopez et al., 2022) could provide the required raw data. The remaining two studies were only included in narrative synthesis but not meta-analysis.

#### Exposures

We included all TBIs with severity ranging from mild (including concussion) to severe. For methods of measurement, validated screening tools (including the Ohio State University TBI Identification Method [OSU TBI-ID]; Corrigan & Bogner, 2007), structured clinical interviews, medical records reviews, and clinical diagnosis were included. Regarding the major exposure data, we primarily extracted the number of participants experiencing TBI.

#### Study characteristics

For the characteristics of included studies, apart from the above exposure and outcome data items, we also extracted the (i) year and location of the study, (ii) study design, and (iii) participant characteristics (in the exposure and control groups, if any).

### Quality assessment

Two reviewers (KCY & GR) independently assessed the quality of included studies using Kmet et al.’s (2004) quality assessment scale. This consisted of a 14-item checklist on a 3-point scale (0 = criteria not met; 1 = partially met; 2 = fully met) generating a summary score (total sum / total possible sum) ranging from 0 to 100, to categorise the low (≤ 54), moderate (55–74), and high (≥ 75) quality of evidence. The areas of assessment included evaluation of appropriateness of research objectives, study design, sampling methods, recruitment of participants, adoption of measures, sample size, statistical analyses, estimate of variance, control for confounders, results reported, and conclusion drawn. All disagreements were resolved by consensus.

### Synthesis methods

We estimated the meta-analytic odds ratio (OR) with 95% confidence interval (CI) of psychotic disorders or symptoms associated with preceding paediatric TBI among the included studies using the *R* package ‘meta’ (Balduzzi, Rücker, & Schwarzer, 2019). We computed the *I²* statistic to measure heterogeneity among included studies, and the levels of low, moderate, and high heterogeneity were assigned to *I²* values of 25%, 50%, and 75% respectively (Higgins et al., 2003). We expected a moderate-to-high *I²* value due to methodological heterogeneity, and subsequently we opted to use a random-effects model to estimate pooled estimates using the Paule–Mandel estimator (Paule & Mandel, 1982) given evidence for its lower risk of bias compared to other methods (Langan, Higgins, & Simmonds, 2017). We used a funnel plot to test for evidence of publication bias and Egger’s test was planned to provide a statistical test of funnel plot asymmetry (Ioannidis & Trikalinos, 2007). Subgroup analyses based on (i) study location, (ii) study design (i.e., case-control study versus cohort study), (iii) type of outcome being measured (i.e. psychotic disorders versus symptoms or sub-threshold symptoms of psychosis), (iv) type of outcome measurement (i.e. clinical diagnosis vs validated/structured method), and (v) time of onset of the outcomes (i.e. childhood/adolescence versus adulthood) were conducted. We followed the suggested guidelines reporting any detections of statistically significant subgroup differences (Richardson, Garner, & Donegan, 2019). Afterwards, we conducted Viechtbauer and Cheung’s (2010)’s outlier and influential study diagnostics and a leave-one-out sensitivity analysis to assess the presence of any overly-influential studies in estimating the pooled effect. We also completed a meta-regression to estimate whether study quality was related to study outcome. If the meta-regression was statistically significant, a sensitivity analysis was performed to assess whether the pooled association remained robust after removing studies of low quality.

All analyses were conducted with *R* (version 4.2.1; R Core Team, 2020) and were conducted on a Linux x86_64 platform. All *R* code and data for the analyses are available online in the following archive: https://github.com/vaughanbell/pTBI_psychosis_meta-analysis

For any studies that did not yield meta-analysed results, we planned to conduct a narrative synthesis to assess how the additional studies might affect the interpretation of the overall findings, using ESRC guidelines (Popay et al., 2006).

## Results

### Study selection

A total of 850 records resulted from searching the PsycINFO (*n* = 365) and MEDLINE (*n* = 485) databases. After removing duplicates by Ovid’s automatic de-duplication feature, 688 records remained. Seventy records were eligible for full-text screening, of these 60 were excluded. A total of 10 studies were included in this review. See figure 1 for PRISMA 2020 flow diagram.

**Figure 1.**
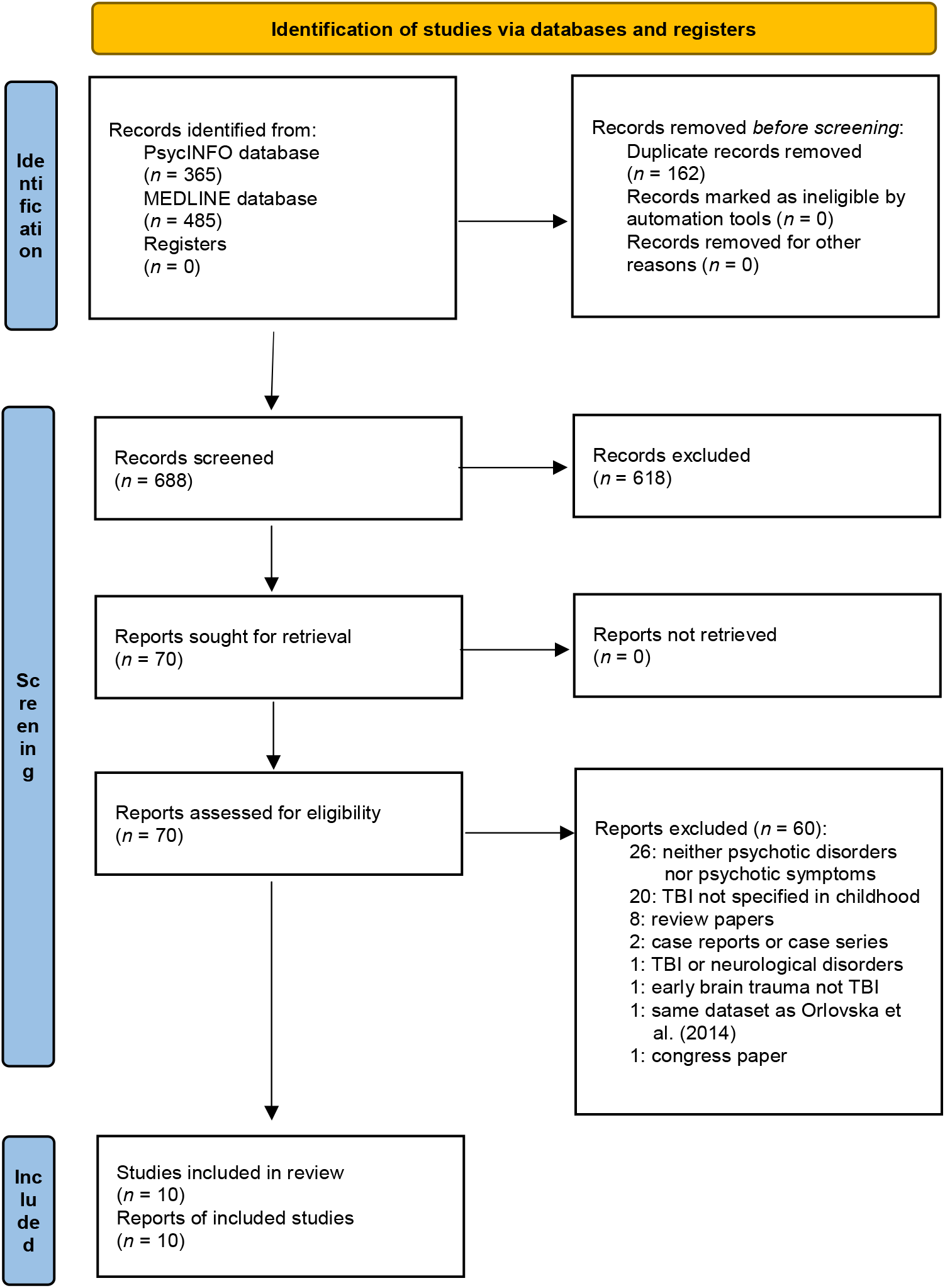
PRISMA 2020 flow diagram for literature search

### Study characteristics

Among the 10 included studies, five adopted case-control designs (AbdelMalik et al., 2003; Deighton et al., 2016; Harrison et al., 2006; Helgeland & Torgersen, 2005; Wilcox & Nasrallah, 1987) and five adopted cohort designs (Ledoux et al., 2022; Lopez et al., 2022; Massagli et al., 2004; Orlovska et al., 2014; Timonen et al., 2002). Four studies were carried out in the United States (Deighton et al., 2016; Lopez et al., 2022; Massagli et al., 2004; Wilcox & Nasrallah, 1987), whilst the remaining six studies were undertaken in other places including Canada (AbdelMalik et al., 2003; Ledoux et al., 2022), Denmark (Orlovska et al., 2014), Finland (Timonen et al., 2002), Norway (Helgeland & Torgersen, 2005), and Sweden (Harrison et al., 2006). Five studies measured schizophrenia as an outcome (AbdelMalik et al., 2003; Harrison et al., 2006; Helgeland & Torgersen, 2005; Timonen et al., 2002; Wilcox & Nasrallah, 1987), four studies measured psychosis (Harrison et al., 2006; Ledoux et al., 2022; Massagli et al., 2004; Orlovska et al., 2014), and two studies investigated sub-threshold symptoms of psychosis (Deighton et al., 2016; Lopez et al., 2022). For the method of outcome measurement, six studies adopted clinical diagnosis (Harrison et al., 2006; Ledoux et al., 2022; Massagli et al., 2004; Orlovska et al., 2014; Timonen et al., 2002; Wilcox & Nasrallah, 1987), whilst the remaining four studies adopted validated psychometric measures or structured clinical interviews (AbdelMalik et al., 2003; Deighton et al., 2016; Helgeland & Torgersen, 2005; Lopez et al., 2022). Finally, in terms of the window of interest regarding the onset of a psychotic disorder or psychotic symptom, six studies reported psychotic disorders or symptoms in adulthood (AbdelMalik et al., 2003; Deighton et al., 2016; Harrison et al., 2006; Orlovska et al., 2014; Timonen et al., 2002; Wilcox & Nasrallah, 1987), whilst the remaining four studies reported childhood and adolescence (Helgeland & Torgersen, 2005; Ledoux et al., 2022; Lopez et al., 2022; Massagli et al., 2004). Detailed characteristics of all the included primary studies are shown in supplementary Table S1. A summary of included study characteristics are presented in Table 2. Details of the quality assessment ratings are reported in supplementary Table S2, with seven studies rated as demonstrating high quality of evidence, one moderate, and two low.

**Table 2.**
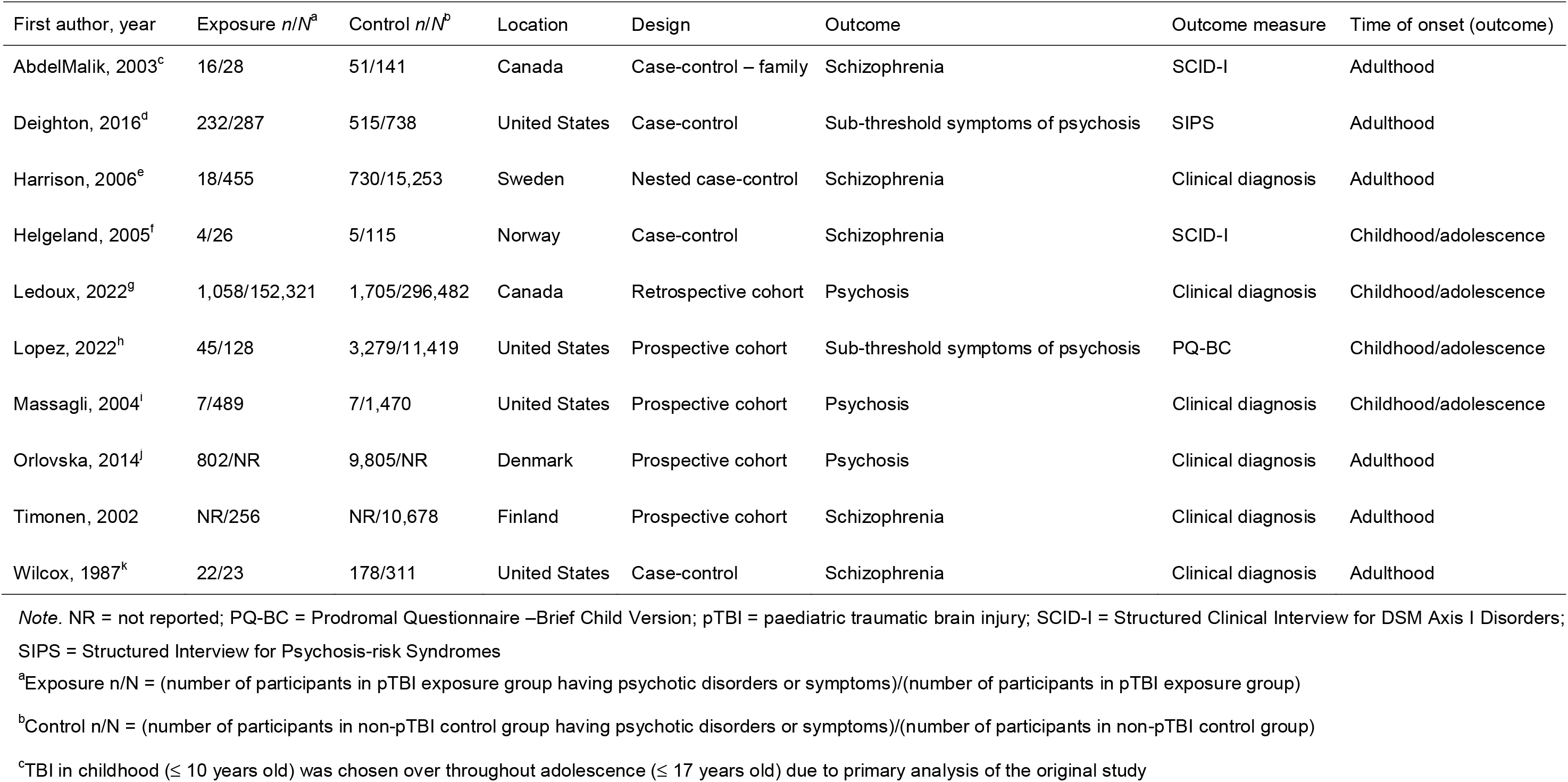
Comparison data for probability of psychotic disorders or symptoms following pTBI.

### Synthesis of results

#### Overall pooled analysis

Among the 10 included studies, raw data from two were either not published or not provided by the original authors after contacts and were therefore excluded from the meta-analysis. Based on eight studies, with a pooled sample size of 479,686 (153,757 in the pTBI group; 325,929 in the control group), there was an overall significant positive association between exposure to paediatric TBI and outcomes of psychotic disorders or symptoms (pooled odds ratio [OR] = 1.80, 95% CI [1.11, 2.95]) with moderate between-study heterogeneity (*I^2^* = 69%, τ^2^ = 0.35, *p* < 0.01). Figure 2 shows the comparison data and forest plot of the corresponding analysis.

**Figure 2.**
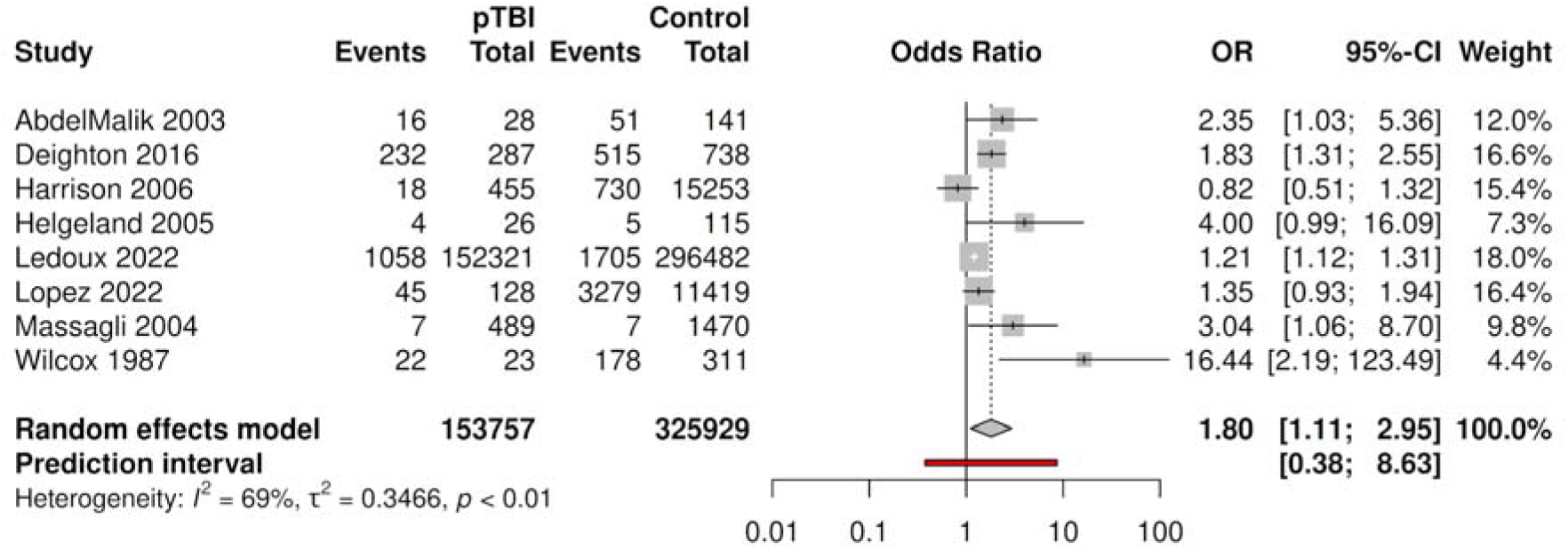
Comparison data and forest plot of odds ratio meta-analysis for psychotic disorders or symptoms

#### Subgroup analyses

Subgroup analyses based on study location (*p* = 0.34), design (*p* = 0.30), psychotic disorder vs subthreshold symptoms of psychosis (*p* = 0.48), measurement type (*p* = 0.77), time of onset (that psychotic disorder/symptoms emerged) (*p* = 0.62) were all non-significant, suggesting that these variables did not modify the effect of paediatric TBI on the probability of psychotic disorders/symptoms. Forest plots of all the above subgroup analyses are reported as Figures S1–S5 in the supplementary material.

In the subgroup analysis comparing psychotic disorder with subthreshold symptoms of psychosis, both remained reliably associated with pTBI (psychotic disorder OR = 2.11, 95% CI [1.01, 4.37]; subthreshold symptoms of psychosis OR = 1.58, 95% CI [1.17, 2.13])

#### Robustness and sensitivity analyses

For the assessment of publication bias, visual inspection of the funnel plot (Figure 3) appeared to exhibit asymmetry. Egger’s test was completed (p = 0.052) although was likely under-powered given 10 studies are considered the minimum for a reliable assessment of publication bias (Ioannidis & Trikalinos, 2007).

**Figure 3.**
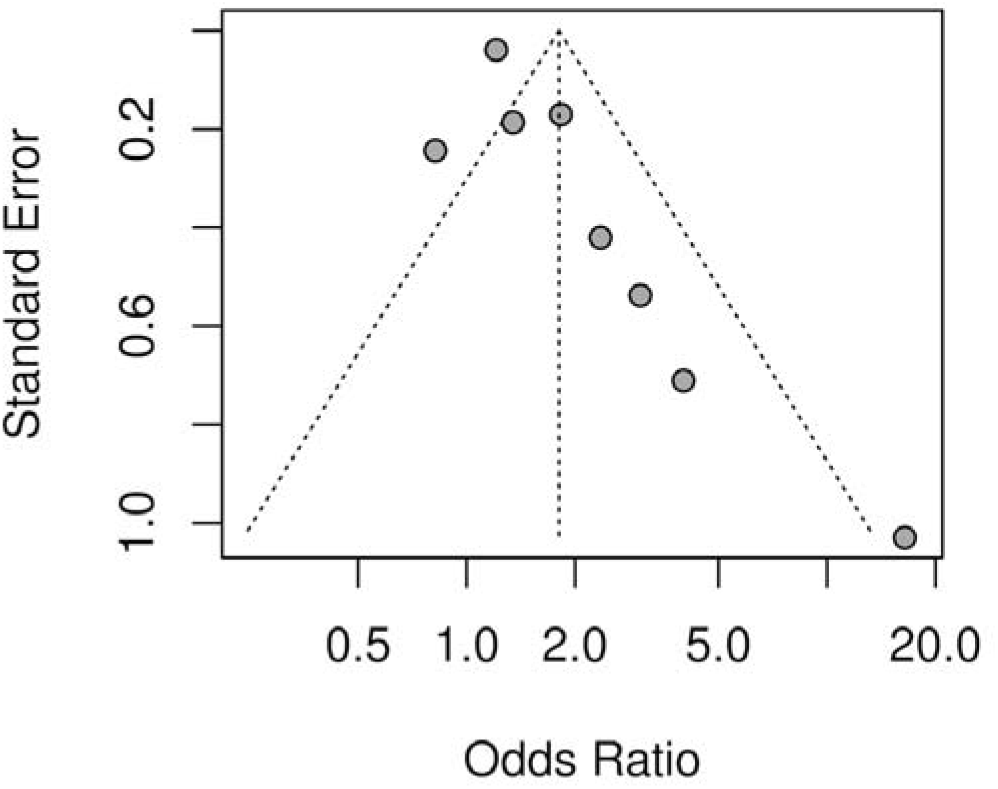
Funnel plot of standard error by odds ratio in meta-analysis

No studies were identified as outliers but one study (Wilcox & Nasrallah, 1987) was identified as excessively influential using Viechtbauer and Cheung’s (2010) outlier and influential diagnostics. A leave-one-out sensitivity analysis showed that removing Wilcox & Nasrallah (1987) reduced the pooled estimate although the association between pTBI and psychosis outcomes remained reliable (OR = 1.52 95% CI [1.08; 2.14]). The removal of one study altered the estimate to the non-significant range, namely Massagli et al. (2004) revised estimate OR = 1.74, 95% CI [0.9997, 3.04]).

A meta-regression analysis indicated that the quality of evidence summary score predicted the association between pTBI and psychotic disorders/symptoms, albeit weakly (random-effects estimate = −0.037, 95% CI [-0.06, −0.01], p = 0.003). Consequently, we completed a sensitivity analysis removing studies with evidence rated as low quality and recalculating the pooled estimate. The revised pooled estimate (see Figure 4) remained significant with narrower confidence intervals suggesting a more accurate estimate (OR = 1.43, 95% CI [1.04, 1.98]) and slightly reduced heterogeneity (*I^2^* = 64%, τ^2^ = 0.10, *p* = 0.02).

**Figure 4.**
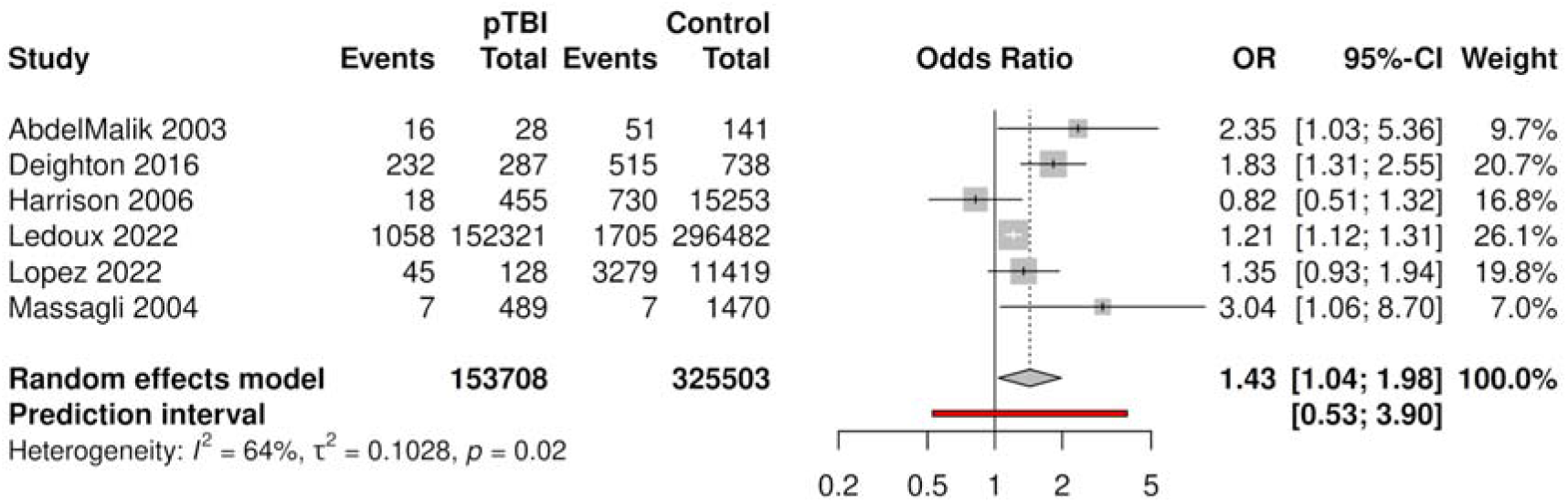
Comparison data and forest plot of odds ratio meta-analysis for psychotic disorders or symptoms – sensitivity analysis by the removal of studies with low quality of evidence

#### Narrative synthesis including additional studies

Two cohort studies were not included in the meta-analysis due to insufficient data, namely Orlovska et al. (2014) and Timonen et al. (2002). Orlovska et al. reported that, compared with individuals without hospital contact for head injury, those exposed to head injury between ages 0 and 5 years had higher rates of schizophrenia spectrum disorders (adjusted incident rate ratio [aIRR] = 1.35, 95% CI [1.18, 1.54]). In addition, differing effects of head injury at 6–10 years (aIRR = 1.33, 95% CI [1.16, 1.50]) and 11–15 years (aIRR = 1.86, 95% CI [1.66, 2.07]) were observed. In Timonen et al, although the original analysis focussed on the association between preceding paediatric TBI and the broad outcomes of mental disorders, Molloy et al. (2011) contacted the original authors and reported no association between paediatric TBI and the subsequent development of schizophrenia (odds ratio [OR] = 1.1, 95% CI [0.41, 2.96]) although the wide confidence intervals indicate that the estimate would carry less weight in estimating an overall effect.

## Discussion

We conducted a systematic review and meta-analysis to estimate the association between paediatric TBI and psychosis, including both frank psychotic disorders and psychotic symptoms. Based on a pooled sample size of 479,686, it was found that pTBI was associated with an increased probability of psychotic disorders and symptoms, with moderate between-study heterogeneity. Regarding the robustness of findings, the estimated association passed robustness tests for study quality, outliers, and excessively influential studies, although the removal of one would have reduced confidence in a reliable association in a leave-one-out sensitivity analysis. This reflects the fact that the lower bound of the confidence interval for the pooled estimate was only marginally above one and therefore confidence in the reliability of this estimate must be taken cautiously. Two studies were identified in the systematic review which could not be included in the meta-analysis, and these studies reporting conflicting results. However, given the characteristics of these additional studies, we consider that including them would have moderately increased our confidence in an association between pTBI and psychosis. Consequently, we conclude that this analysis provides additional evidence for an association between pTBI and psychosis. However, concerns remain about the long-run reliability of this estimate remain and new studies will be crucial in deciding this issue.

In relating the above findings to the field, our results raise the feasibility of potential causal associations between pTBI and psychosis. Developmental models of psychosis suggest that pTBI could be a plausible risk factor for psychosis, given that the full spectrum of pTBI (from mild pTBI to severe brain injury) has established effects on neurodevelopment (Emery et al., 2016; Goh et al., 2021), and events that have an adverse impact on neurodevelopment are known risk factors for psychosis (Howes & Murray, 2014; Murray et al., 2017). Our meta-analytic results seem to suggest the role of pTBI as a risk factor for psychosis. However, reverse causality or shared risk factor pathways are also possible. Brain injuries have been hypothesised to be more common in young people who have a higher risk for psychosis, as they may already show subtle premorbid difficulties such as motor coordination leading to a higher risk for accidental injuries (AbdelMalik et al., 2003; David & Prince, 2005). Furthermore, psychotic symptoms, including in people without frank psychosis, are associated with higher rates of early-life bullying (Catone et al., 2015; Valmaggia et al., 2015), suggesting a possible reverse or reciprocal association between psychotic spectrum phenomena and acquired brain injury through victimisation violence. A well-designed prospective cohort study would be needed to reliably identify relationships between pTBI, psychotic symptoms, and any potential confounders and/or mediators.

Nevertheless, given the concordance between the findings reported here and developmental models of psychosis (Howes & Murray, 2014; Murray et al., 2017), this may suggest that history of pTBI may be a useful addition when taking a history of patients with psychosis and that public health that prevent pTBI may have longer-term benefits for lifetime mental health.

A strength of this study is the use of systematic procedures to comprehensively search for eligible studies. We also included sensitivity analyses to ensure robustness of the findings. In addition, we pre-registered the review to reduce the risk of bias. Moreover, the meta-analysis is available online as a computational notebook with an open dataset, to enhance openness and reproducibility.

However, we note several limitations of this study. The first is that the included studies are heterogeneous in terms of their measured outcomes design (case control vs cohort), outcome (symptoms vs disorder), outcome measure (clinical diagnosis vs validated measure) and life-stage of measured psychosis outcome (adulthood vs childhood/adolescence). Our subgroup analyses found no evidence for difference of association between subgroups. We note the potential for low statistical power to make identifying associations within subgroups difficult, given than subgroups typically included 3-4 studies. However, the heterogeneity of studies reflects the fact that many were not primarily designed to assess the association with pTBI and psychosis spectrum phenomena, and more focused and better design studies are clearly needed. Inspection of the funnel plot indicated a potential for publication bias which could have reduced the accuracy or the direction of the estimate. In addition, studies either did not report severity of brain injury, or did not distinguish between severity, meaning it was not possible to examine whether there was a ‘dose-response’ relationship between TBI and later psychosis, a potentially important consideration when examining evidence for causality. We also note that we solely included studies published in English, and listed in primarily English language databases, potentially missing some potentially useful evidence.

Based on the above discussions, it is recommended that future research focus on specifically assessing the association between pTBI and psychosis spectrum phenomena. In addition, to rule out the possible reverse association, a well-designed prospective cohort study would be needed to reliably identify relationships between pTBI, psychotic symptoms, and any potential confounders and/or mediators. Additionally, future studies specifying the type of TBI (e.g. due to accidents or other sources), and location and type of injury, would enhance our understanding of the presence/absence of an aetiological relationship between pTBI and psychosis. Lastly, future reviews should consider including non-English language databases.

In conclusion, our systematic review and meta-analysis reports evidence for a positive association between paediatric TBI and subsequent outcomes of psychotic disorders or symptoms but with caveats regarding our confidence in the long-term reliability of this association as new evidence emerges.

## Data Availability

All data produced are available online at https://github.com/vaughanbell/pTBI_psychosis_meta-analysis

## References

AbdelMalik, P., Husted, J., Chow, E. W. C., & Bassett, A. S. (2003). Childhood Head Injury and Expression of Schizophrenia in Multiply Affected Families. Archives of General Psychiatry, 60(3), 231–236. https://doi.org/10.1001/archpsyc.60.3.231

Balduzzi, S., Rücker, G., & Schwarzer, G. (2019). How to perform a meta-analysis with R: a practical tutorial. Evidence-Based Mental Health, 22(4), 153–160.

Batty, R. A., Rossell, S. L., Francis, A. J. P., & Ponsford, J. (2013). Psychosis Following Traumatic Brain Injury. Brain Impairment, 14(1), 21–41. https://doi.org/10.1017/BrImp.2013.10

Carroll, L. J., Cassidy, J. D., Cancelliere, C., Côté, P., Hincapié, C. A., Kristman, V. L., … Hartvigsen, J. (2014). Systematic Review of the Prognosis After Mild Traumatic Brain Injury in Adults: Cognitive, Psychiatric, and Mortality Outcomes: Results of the International Collaboration on Mild Traumatic Brain Injury Prognosis. Archives of Physical Medicine and Rehabilitation, 95(3, Supplement), S152–S173. https://doi.org/10.1016/j.apmr.2013.08.300

Catone, G., Marwaha, S., Kuipers, E., Lennox, B., Freeman, D., Bebbington, P., & Broome, M. (2015). Bullying victimisation and risk of psychotic phenomena: Analyses of British national survey data. The Lancet Psychiatry, 2(7), 618–624. https://doi.org/10.1016/S2215-0366(15)00055-3

Cnossen, M. C., Scholten, A. C., Lingsma, H. F., Synnot, A., Haagsma, J., Steyerberg, Prof. E. W., & Polinder, S. (2017). Predictors of Major Depression and Posttraumatic Stress Disorder Following Traumatic Brain Injury: A Systematic Review and Meta-Analysis. The Journal of Neuropsychiatry and Clinical Neurosciences, 29(3), 206–224. https://doi.org/10.1176/appi.neuropsych.16090165

Corrigan, J. D., & Bogner, J. (2007). Initial Reliability and Validity of the Ohio State University TBI Identification Method. The Journal of Head Trauma Rehabilitation, 22(6), 318–329. https://doi.org/10.1097/01.HTR.0000300227.67748.77

David, A. S., & Prince, M. (2005). Psychosis following head injury: A critical review. Journal of Neurology, Neurosurgery & Psychiatry, 76(suppl 1), i53–i60. https://doi.org/10.1136/jnnp.2004.060475

Deighton, S., Buchy, L., Cadenhead, K. S., Cannon, T. D., Cornblatt, B. A., McGlashan, T. H., … Addington, J. (2016). Traumatic brain injury in individuals at clinical high risk for psychosis. Schizophrenia Research, 174(1), 77–81. https://doi.org/10.1016/j.schres.2016.04.041

Emery, C. A., Barlow, K. M., Brooks, B. L., Max, J. E., Villavicencio-Requis, A., Gnanakumar, V., … Yeates, K. O. (2016). A Systematic Review of Psychiatric, Psychological, and Behavioural Outcomes following Mild Traumatic Brain Injury in Children and Adolescents. The Canadian Journal of Psychiatry, 61(5), 259–269. https://doi.org/10.1177/0706743716643741

Feighner, J. P., Robins, E., Guze, S. B., Woodruff, R. A., Jr., Winokur, G., & Munoz, R. (1972). Diagnostic Criteria for Use in Psychiatric Research. Archives of General Psychiatry, 26(1), 57–63. https://doi.org/10.1001/archpsyc.1972.01750190059011

Fleminger, S. (2008). Long-term psychiatric disorders after traumatic brain injury. European Journal of Anaesthesiology | EJA, 25, 123. https://doi.org/10.1017/S0265021507003250

Fujii, D., & Ahmed, I. (2002). Characteristics of Psychotic Disorder Due to Traumatic Brain Injury. The Journal of Neuropsychiatry and Clinical Neurosciences, 14(2), 130–140. https://doi.org/10.1176/jnp.14.2.130

Goh, M. S. L., Looi, D. S. H., Goh, J. L., Sultana, R., Goh, S. S. M., Lee, J. H., & Chong, S.-L. (2021). The Impact of Traumatic Brain Injury on Neurocognitive Outcomes in Children: A Systematic Review and Meta-Analysis. Journal of Neurology, Neurosurgery & Psychiatry, 92(8), 847–853. https://doi.org/10.1136/jnnp-2020-325066

Harrison, G., Whitley, E., Rasmussen, F., Lewis, G., Dalman, C., & Gunnell, D. (2006). Risk of schizophrenia and other non-affective psychosis among individuals exposed to head injury: Case control study. Schizophrenia Research, 88(1), 119–126. https://doi.org/10.1016/j.schres.2006.07.001

Helgeland, M. I., & Torgersen, S. (2005). Stability and prediction of schizophrenia from adolescence to adulthood. European Child & AdolescentPsychiatry, 14(2), 83–94. https://doi.org/10.1007/s00787-005-0436-0

Howes, O. D., & Murray, R. M. (2014). Schizophrenia: An integrated sociodevelopmental-cognitive model. The Lancet, 383(9929), 1677–1687. https://doi.org/10.1016/S0140-6736(13)62036-X

Ioannidis, J. P. A., & Trikalinos, T. A. (2007). The appropriateness of asymmetry tests for publication bias in meta-analyses: A large survey. CMAJLJ: Canadian Medical Association Journal, 176(8), 1091–1096. https://doi.org/10.1503/cmaj.060410

Karcher, N. R., Barch, D. M., Avenevoli, S., Savill, M., Huber, R. S., Simon, T. J., … Loewy, R. L. (2018). Assessment of the Prodromal Questionnaire–Brief Child Version for Measurement of Self-reported Psychoticlike Experiences in Childhood. JAMA Psychiatry, 75(8), 853–861. https://doi.org/10.1001/jamapsychiatry.2018.1334

Kmet, L. M., Cook, L. S., & Lee, R. C. (2004). Standard quality assessment criteria for evaluating primary research papers from a variety of fields.

Langan, D., Higgins, J. P. T., & Simmonds, M. (2017). Comparative performance of heterogeneity variance estimators in meta-analysis: A review of simulation studies. Research Synthesis Methods, 8(2), 181–198. https://doi.org/10.1002/jrsm.1198

Ledoux, A.-A., Webster, R. J., Clarke, A. E., Fell, D. B., Knight, B. D., Gardner, W., … Zemek, R. (2022). Risk of Mental Health Problems in Children and Youths Following Concussion. JAMA Network Open, 5(3), e221235. https://doi.org/10.1001/jamanetworkopen.2022.1235

Lee, K.-W., Chan, K.-W., Chang, W.-C., Lee, E. H.-M., Hui, C. L.-M., & Chen, E. Y.-H. (2016). A systematic review on definitions and assessments of psychotic-like experiences. Early Intervention in Psychiatry, 10(1), 3–16. https://doi.org/10.1111/eip.12228

Linscott, R. J., & van Os, J. (2013). An updated and conservative systematic review and meta-analysis of epidemiological evidence on psychotic experiences in children and adults: On the pathway from proneness to persistence to dimensional expression across mental disorders. Psychological Medicine, 43(6), 1133–1149. https://doi.org/10.1017/S0033291712001626

Lopez, D. A., Christensen, Z. P., Foxe, J. J., Ziemer, L. R., Nicklas, P. R., & Freedman, E. G. (2022). Association between mild traumatic brain injury, brain structure, and mental health outcomes in the Adolescent Brain Cognitive Development Study. NeuroImage, 263, 119626. https://doi.org/10.1016/j.neuroimage.2022.119626

Massagli, T. L., Fann, J. R., Burington, B. E., Jaffe, K. M., Katon, W. J., & Thompson, R. S. (2004). Psychiatric illness after mild traumatic brain injury in children11No commercial party having a direct financial interest in the results of the research supporting this article has or will confer a benefit on the author(s) or on any organization with which the author(s) is/are associated. Archives of Physical Medicine and Rehabilitation, 85(9), 1428–1434. https://doi.org/10.1016/j.apmr.2003.12.036

McGlashan, T., Walsh, B., & Woods, S. (2010). The psychosis-risk syndrome: Handbook for diagnosis and follow-up. Oxford University Press.

Molloy, C., Conroy, R. M., Cotter, D. R., & Cannon, M. (2011). Is Traumatic Brain Injury A Risk Factor for Schizophrenia? A Meta-Analysis of Case-Controlled Population-Based Studies. Schizophrenia Bulletin, 37(6), 1104–1110. https://doi.org/10.1093/schbul/sbr091

Murray, R. M., Bhavsar, V., Tripoli, G., & Howes, O. (2017). 30 Years on: How the Neurodevelopmental Hypothesis of Schizophrenia Morphed Into the Developmental Risk Factor Model of Psychosis. Schizophrenia Bulletin, 43(6), 1190–1196. https://doi.org/10.1093/schbul/sbx121

Orlovska, S., Pedersen, M. S., Benros, M. E., Mortensen, P. B., Agerbo, E., & Nordentoft, M. (2014). Head Injury as Risk Factor for Psychiatric Disorders: A Nationwide Register-Based Follow-Up Study of 113,906 Persons With Head Injury. American Journal of Psychiatry, 171(4), 463–469. https://doi.org/10.1176/appi.ajp.2013.13020190

Os, J. van, Linscott, R. J., Myin-Germeys, I., Delespaul, P., & Krabbendam, L. (2009). A systematic review and meta-analysis of the psychosis continuum: Evidence for a psychosis proneness–persistence–impairment model of psychotic disorder. Psychological Medicine, 39(2), 179–195. https://doi.org/10.1017/S0033291708003814

Page, M. J., McKenzie, J. E., Bossuyt, P. M., Boutron, I., Hoffmann, T. C., Mulrow, C. D., … Moher, D. (2021). The PRISMA 2020 statement: An updated guideline for reporting systematic reviews. BMJ, 372, n71. https://doi.org/10.1136/bmj.n71

Paule, R. C., & Mandel, J. (1982). Consensus values and weighting factors. Journal of Research of the National Bureau of Standards, 87(5), 377.

Perry, D. C., Sturm, V. E., Peterson, M. J., Pieper, C. F., Bullock, T., Boeve, B. F., … Welsh-Bohmer, K. A. (2016). Association of traumatic brain injury with subsequent neurological and psychiatric disease: A meta-analysis. Journal of Neurosurgery, 124(2), 511–526. https://doi.org/10.3171/2015.2.JNS14503

Popay, J., Roberts, H., Sowden, A., Petticrew, M., Arai, L., Rodgers, M., … Duffy, S. (2006). Guidance on the conduct of narrative synthesis in systematic reviews. A Product from the ESRC Methods Programme Version, 1(1), b92.

R Core Team. (2020). R: A language and environment for statistical computing. Vienna, Austria: R Foundation for Statistical Computing. Retrieved from https://www.R-project.org/

Richardson, M., Garner, P., & Donegan, S. (2019). Interpretation of subgroup analyses in systematic reviews: A tutorial. Clinical Epidemiology and Global Health, 7(2), 192–198. https://doi.org/10.1016/j.cegh.2018.05.005

Rogers, J. M., & Read, C. A. (2007). Psychiatric comorbidity following traumatic brain injury. Brain Injury, 21(13–14), 1321–1333. https://doi.org/10.1080/02699050701765700

Rubio, J. M., Sanjuan, J., Florez-Salamanca, L., & Cuesta, M. J. (2012). Examining the course of hallucinatory experiences in children and adolescents: A systematic review. SCHIZOPHRENIA RESEARCH, 138(2–3), 248–254. https://doi.org/10.1016/j.schres.2012.03.012

Schwartz, J. A., Jodis, C. A., Breen, K. M., & Parker, B. N. (2019). Brain injury and adverse outcomes: A contemporary review of the evidence. Current Opinion in Psychology, 27, 67–71. https://doi.org/10.1016/j.copsyc.2018.09.006

Timonen, M., Miettunen, J., Hakko, H., Zitting, P., Veijola, J., von Wendt, L., & Räsänen, P. (2002). The association of preceding traumatic brain injury with mental disorders, alcoholism and criminality: The Northern Finland 1966 Birth Cohort Study. Psychiatry Research, 113(3), 217–226. https://doi.org/10.1016/S0165-1781(02)00269-X

Trotta, A., Murray, R. M., & Fisher, H. L. (2015). The impact of childhood adversity on the persistence of psychotic symptoms: A systematic review and meta-analysis. Psychological Medicine, 45(12), 2481–2498. https://doi.org/10.1017/S0033291715000574

Valmaggia, L. R., Day, F. L., Kroll, J., Laing, J., Byrne, M., Fusar-Poli, P., & McGuire, P. (2015). Bullying victimisation and paranoid ideation in people at ultra high risk for psychosis. Schizophrenia Research, 168(1), 68–73. https://doi.org/10.1016/j.schres.2015.08.029

van Reekum, R., Cohen, T., & Wong, J. (2000). Can Traumatic Brain Injury Cause Psychiatric Disorders? The Journal of Neuropsychiatry and Clinical Neurosciences, 12(3), 316–327. https://doi.org/10.1176/jnp.12.3.316

Viechtbauer, W., & Cheung, M. W.-L. (2010). Outlier and influence diagnostics for meta-analysis. Research Synthesis Methods, 1(2), 112–125. https://doi.org/10.1002/jrsm.11

Wilcox, J. A., & Nasrallah, H. A. (1987). Childhood head trauma and psychosis. Psychiatry Research, 21(4), 303–306. https://doi.org/10.1016/0165-1781(87)90013-8

