## Supplementary material for "Paediatric traumatic brain injury as a risk factor for psychosis and psychotic symptoms: a systematic review and meta-analysis"

**Appendix S1**

PsycINFO (from 1806) and MEDLINE (from 1946) databases were searched via Ovid.

1. Brain Injuries/

2. Traumatic Brain Injury/

3. Brain Concussion/

4. Psychosis/

5. Schizophrenia/

6. (TBI or traumatic brain injur* or brain injur* or head injur* or cerebral trauma or craniocerebral injur* or concussion* or skull fracture*).ab,id,ti.

7. (psychosis or psychotic or psychotic disorder* or psychotic exp* or psychotic?like exp* or schizophreni* or delusional disorder* or delusion* or hallucinat* or psychiatric illness* or psychiatric disorder*).ab,id,ti.

8. (infan* or baby* or babies or toddler* or preschool* or child* or pediat* or paediat* or prepubescen* or prepuberty* or puberty or pubescen* or teen* or young* or youth* or minors* or underag* or juvenile* or preadolesc* or adolesc*).ab,id,ti.

9. (childhood birth 12 yrs or preschool age 2 5 yrs or school age 6 12 yrs or adolescence 13 17 yrs).ag.

10. 1 or 2 or 3 or 6

11. 4 or 5 or 7

12. 8 or 9

13. 10 and 11 and 12

14. Brain Injuries/

15. Brain Injuries, Traumatic/

16. Brain Concussion/

17. Psychotic Disorders/

18. Schizophrenia/

19. Infant/

20. Child, Preschool/

21. Child/

22. Adolescent/

23. (TBI or traumatic brain injur* or brain injur* or head injur* or cerebral trauma or craniocerebral injur* or concussion* or skull fracture*).ab,kw,ti.

24. (psychosis or psychotic or psychotic disorder* or psychotic exp* or psychotic?like exp* or schizophreni* or delusional disorder* or delusion* or hallucinat* or psychiatric illness* or psychiatric disorder*).ab,kw,ti.

25. (infan* or baby* or babies or toddler* or preschool* or child* or pediat* or paediat* or prepubescen* or prepuberty* or puberty or pubescen* or teen* or young* or youth* or minors* or underag* or juvenile* or preadolesc* or adolesc*).ab,kw,ti.

26. 14 or 15 or 16 or 23

27. 17 or 18 or 24

28. 19 or 20 or 21 or 22 or 25

29. 26 and 27 and 28

30. 13 or 29

31. remove duplicates from 30

**Table S1**. Characteristics of primary studies from MEDLINE & PsycINFO included in systematic review

| Study, year  (location) | Study design | Participant | Exposure | Outcome | Finding | Sig. assoc. |
| --- | --- | --- | --- | --- | --- | --- |
| AbdelMalik  2003  (Canada) | Case-control – family | 169 individuals (67 with narrowly defined schizophrenia; 102 siblings without schizophrenia) from 23 Canadian families with schizophrenia | Modified Structured Clinical Interview for DSM-III-R (SCID-I), supplemented by collateral information from medical records and family. Occurrence & severity of childhood head injuries (**≤** 10 years old) rated by three investigators independently. TBI severity ranging from possible mild to mild. | SCID-I by a psychiatrist to diagnose narrowly defined schizophrenia (i.e., schizophrenia or chronic schizoaffective disorder) | Participants in the schizophrenia group (*n* = 16 [23.9%]) had a higher likelihood than the unaffected siblings group (*n* = 12 [11.8%]) to have a history of head injury in childhood (OR = 2.35, 95% CI [1.03, 5.36], *p* = 0.04) | + |
| Deighton  2016  (United States) | Case-control | 1,025 help-seekers (747 clinical high risk [CHR] of psychosis; 278 healthy controls [HC]) recruited from the 8-site North American Prodrome Longitudinal Study (NAPLS 2) | Traumatic Brain Injury (TBI) Interview, assessing previous history of TBI, including the age at first TBI, age at most recent TBI, count of the number of TBIs, and severity of TBI. Only mild TBI was included | Structured Interview for Psychosis-risk Syndromes (SIPS) to assess the Criteria of Psychosis-risk Syndromes (COPS). If the Presence of Psychotic Symptoms Criteria (POPS) was met, further clinical assessment to determine diagnosis of psychosis | Participants in the CHR group experienced a mild TBI (*n* = 232 [31.0%]) more often than the HC (*n* = 55 [19.8%]) (χ^2^ = 12.77, *p* < 0.001)  CHR participants who experienced a mild TBI and later made the transition to psychosis were significantly younger at the age at first (*M* = 7.8, *SD* = 3.0) and most recent TBI (*M* = 10.1, *SD* = 5.5), than those who did not develop psychosis (*M* of age at first TBI = 10.6, *SD* = 5.6, U = 1732.00, *p* = 0.02) (*M* of age at most recent TBI = 12.4, *SD* = 6.0, U = 1818.50, *p* = 0.04) | + |
| Harrison  2006  (Sweden) | Nested case-control | Swedish individuals born between 1973 and 1980 (748 cases of schizophrenia and 14,960 matched controls; 1,526 non-affective psychosis and 30,520 matched controls) from a cohort of 731,305 members obtained from several linked Swedish registers | Swedish Inpatient Discharge Register to identify hospital admission for concussion with/without any face/head/skull injuries, as well as all skull/intracranial injuries (ICD-10 codes: S02.0, S02.1, S02.7–S02.9, S06.0, S06.1–S06.9, S09.7). Only severe head injury was included. Timing of the head injury was collected to determine childhood exposure (< 10 years old) | Swedish Inpatient Discharge Register to identify cases of schizophrenia (ICD-10 code: F20) and non-affective psychosis (ICD-10 codes: F21-29) | Participants in the non-affective psychosis group (*n* = 131 [8.6%]) were more likely than the matched control group (*n* = 1,918 [6.3%]) to have a history of severe head injury (aOR = 1.37, 95% CI [1.14, 1.66], *p* = 0.001), but no association was found when comparing the schizophrenia group (*n* = 54 [7.2%]) and matched controls (*n* = 986 [6.6%]) for the likelihood of previous severe head injury (aOR = 1.10, 95% CI [0.82 1.47], *p* = 0.51)  No evidence of the effects of childhood exposure to head injury (< 10 years old) on non-affective psychosis (aOR = 0.94, 95% CI [0.68, 1.29], *p* = 0.70) or schizophrenia (aOR = 0.81, 95% CI [0.50, 1.31], *p* = 0.38) was found | – |
| Helgeland  2005  (Norway) | Case-control | 145 patients (13 cases of schizophrenia; 132 controls without schizophrenia) admitted to the adolescent unit at The National Centre for Child and Adolescent Psychiatry (NCCAP) in Norway from 1963 to 1978 | Hospital records of concussion and head traumas. Further blind review of the detailed medical records by the first author. TBI severity not reported. | Hospital records of psychiatric diagnoses. The detailed records were anonymised in advance by second author, and reviewed by first author with blinding based on the Structured Clinical Interview for DSM-IV Axis I Disorders (SCID-I), to ascertain diagnosis of schizophrenia (early onset was defined as < 18 years old) | Participants with an early onset of schizophrenia (< 18 years old) (*n* = 3 [33.3%]) were significantly more likely than the control group without schizophrenia (*n* = 12 [9.1%]) to have a history of head traumas (*p* < 0.05). No evidence was found in the case of concussion (*p* = ns) | +/– |
| Ledoux  2022  (Canada) | Retrospective cohort | 448,803 children and youth aged 5 to 18 years old (152,321 with exposure to concussion; 296,482 controls with orthopaedic injury) presented to an emergency department, primary health care, or mental health practitioner from 2010 to 2020 in Canada | Canadian national healthcare databases capturing data on visits to emergency department (ICD-10-CA codes: S06.0) and primary health care (Ontario Health Insurance Plan [OHIP] diagnosis code: 850) to ascertain concussion. TBI severity was mild. | National healthcare databases capturing data on psychiatric hospitalisation (ICD-10-CA codes: F20, F22, F23, F24, F25, F28, F29; OHIP codes: 295, 297, 298; and Ontario Mental Health Reporting System [OMHRS] codes: Q1E_RETIRED_2016  Q1B) to establish schizophrenia | The exposed group had a higher risk of subsequently developing mental health conditions (including other non-schizophrenia diagnoses) when compared with the non-exposed control group (aHR = 1.39, 95% CI [1.37, 1.40], *p* < 0.001)  Data on the development of schizophrenia in the exposed (*n* = 1,058 [0.7%]) and non-exposed control group (*n* = 1,705 [0.6%]) were reported in the supplemental materials | NR |
| Lopez  2022  (United States) | Prospective cohort | 11,876 children aged 9-10 (128 with exposure to mild TBI; 322 possible mild TBI; 11,415 controls without TBIs) from 21 research sites at the United States from the Adolescent Brain Cognitive Development (ABCD) Study, with year-1 and year-2 follow-up | The Ohio State University TBI Identification Method (OSU TBI-ID) – Short Modified, with questions directed at parents or guardians, to assess the number of possible mild and mild TBIs | Prodromal Questionnaire –Brief Child Version (PQ-BC), a self-report instrument for children and adolescents, to measure the number of distressing psychotic-like experiences (PLEs) in the past month weighted by level of distress | The exposed-group children with mild TBI had a non-significant 22% increased risk of experiencing distressing PLEs (aIRR = 1.22, 95% CI [0.94, 1.57], *p* = 0.1395) when compared with the non-exposed control group, using imputed data | – |
| Massagli  2004  (United States) | Prospective cohort | 1,960 children (**≤** 14 years old; 490 sustained a mild TBI; 1,470 matched controls) attending emergency department, hospital, or outpatient clinic in the Washington State in 1993, with a 3-year follow-up | Washington State’s counties’ healthcare database capturing data on visits to emergency department, hospitals, or outpatient clinics, to indicate mild TBI including skull fractures (ICD-9-CM codes: 800.0–801.9, 803.0–804.9) and intracranial injury such as concussion (ICD-9-CM codes: 850.0–854.1). Mild TBI was indicated by less than 1-hour or no loss of consciousness and no traumatic intracranial lesions | Washington State’s counties’ healthcare database capturing data on psychiatric diagnosis, prescription for psychiatric medication, or using psychiatric services, to indicate psychotic disorders including organic psychotic disorders (ICD-9-CM codes: 290.0–.9, 293.0–294.9); schizophrenia, hallucinations, paranoia (ICD-9-CM codes: 295, 297.0–299.9, 780.1), and prescription for antipsychotics | The TBI-exposed group children had higher cumulative incidence estimates for any psychiatric illnesses (including psychotic disorders and conditions other than psychotic disorders) in the 3 years (*n* = 146 [30%]) when compared with the non-exposed controls (*n* = 293 [20%]) (*p* = .0001)  The development of psychotic disorder in the TBI-exposed group (*n* = 7 [1.43%]) and non-exposed control group (*n* = 7 [0.48%]) were reported in Table 3 of the original study | NR |
| Orlovska  2014  (Denmark) | Prospective cohort | 1,438,339 individuals born in Denmark between 1977 and 2000 (113,906 with hospital contacts for head injury; 1,324,433 without) included in the Danish nationwide population-based registers, followed for 34 years from 1977 to 2010 | Danish National Hospital Register capturing data on visits to emergency department, inpatient and outpatient services, to identify mild head injury (ICD-10 code: S06.0), skull fracture (ICD-10 codes: S02.0, S02.1, S02.7, S02.9), and severe head injury (ICD-10 codes: S06.1–S06.9) | Danish Psychiatric Central Register capturing data on visits to emergency department, inpatient and outpatient psychiatric services, to identify schizophrenia spectrum disorder (ICD-10 codes: F20-F29) | When compared with those without hospital contact for head injury (*n* = 9,303), the group exposed to head injury between ages 0 and 5 years (*n* = 226) significantly predicted subsequent development of schizophrenia spectrum disorders (aIRR = 1.35, 95% CI [1.18, 1.54]). Significant effects of head injury started from 6–10 years (*n* = 242; aIRR = 1.33, 95% CI [1.16, 1.50]) and 11–15 years (*n* = 334; aIRR = 1.86, 95% CI [1.66, 2.07]) were observed | + |
| Timonen  2002  (Finland) | Prospective cohort | 10,934 individuals (256 with preceding TBI up to age 15 years old; 10,678 without) from the database of the 1966 Birth Cohort Study of Northern Finland, followed through the pre-natal stages prospectively up to the age of 31 years old | Finnish Hospital Discharge Registers capturing data on treatment episodes in hospitals and inpatient wards of health centres nation-wide, to identify TBI (ICD-9 codes: 800–801, 803, 804 except for facial traumas, 850–854, 950–951) up to 15 years old. Case notes of the cohort members with TBI up to 15 years old were screened further by the authors. TBI severity was not reported. | Finnish Hospital Discharge Registers capturing data on treatment episodes in hospitals and inpatient wards of health centres nation-wide, to identify psychiatric disorders (ICD-9 codes for schizophrenia not specified). Case notes of the cohort members with psychiatric disorders were checked against the criteria from DSM-III-R by the authors | The exposure to TBI during childhood and adolescence significantly increased the likelihood of developing mental disorders (aOR = 2.1, 95% CI [1.2, 3.6]) in the male cohort  Although not originally reported by Timonen et al., Molloy et al. (2011) contacted the original authors and reported the following risk estimate for schizophrenia following paediatric TBI (OR = 1.1, 95% CI [0.41, 2.96]) for this study | – |
| Wilcox  1987  (United States) | Case-control | 659 hospitalised patients (200 with schizophrenia; 122 bipolar disorder; 203 depressive disorder; and 134 surgical controls) admitted to a large university hospital from 1934 to 1944 | Hospital records of head traumas. Further blind rating of head injury by the authors without knowledge of psychiatric diagnosis whilst reviewing the medical records  Exposure to childhood head trauma was defined as the onset before 10 years old. TBI severity was not reported. | Diagnosis of schizophrenia based on diagnostic criteria by Feighner et al. (1972) | The group of patients with schizophrenia had significantly more cases of childhood head trauma (*n* = 22 [11%]) when compared with the surgical control group (*n* = 1 [0.7%]) (*p* = 0.0001) and depression group (*n* = 3 [1.5%]) (*p* = 0.0001), but not the bipolar group (*n* = 6 [4.9%]) (*p* = 0.06). | +/– |

*Note.* aHR = adjusted hazard ratio; aIRR = adjusted incidence rate ratio; aOR = adjusted odds ratio; DSM = Diagnostic and Statistical Manual of Mental Disorders; ICD = International Statistical Classification of Diseases and Related Health Problems; NR = not reported; sig. assoc. = significant association; TBI = traumatic brain injury; (+) = significant association; (–) = non-significant association; (+/–) = mixed findings

**Table S2**. Quality assessment ratings for included studies

| Study, year | Checklist for quality assessment^a^ | | | | | | | | | | | Overall quality of evidence (summary score) |
| --- | --- | --- | --- | --- | --- | --- | --- | --- | --- | --- | --- | --- |
|  | Q1: Objective described | Q2:  Appropriate design | Q3:  Appropriate sampling | Q4:  Participant described | Q8:  Well-defined measure | Q9:  Appropriate sample size | Q10:  Appropriate analysis | Q11:  Estimate of variance | Q12:  Confounder controlled | Q13:  Result in detail | Q14:  Conclusion supported |  |
| AbdelMalik, 2003 | ++ | ++ | ++ | ++ | ++ | + | ++ | ++ | + | ++ | ++ | High (90.9) |
| Deighton, 2016 | ++ | ++ | ++ | ++ | ++ | + | ++ | + | + | ++ | ++ | High (86.4) |
| Harrison, 2006 | ++ | ++ | ++ | ++ | + | ++ | ++ | ++ | ++ | ++ | + | High (90.9) |
| Helgeland, 2005 | ++ | + | + | ++ | + | – | + | – | + | + | + | Low (50) |
| Ledoux, 2022 | ++ | ++ | ++ | ++ | + | ++ | ++ | ++ | ++ | ++ | ++ | High (95.5) |
| Lopez, 2022 | ++ | ++ | ++ | ++ | + | ++ | ++ | ++ | + | ++ | ++ | High (90.9) |
| Massagli, 2004 | ++ | ++ | ++ | ++ | + | + | ++ | ++ | + | ++ | ++ | High (86.4) |
| Orlovska, 2014 | ++ | ++ | ++ | + | + | ++ | ++ | ++ | ++ | ++ | ++ | High (90.9) |
| Timonen, 2002 | ++ | ++ | + | + | + | ++ | + | ++ | ++ | + | + | Moderate (72.7) |
| Wilcox, 1987 | + | + | + | – | + | – | + | – | + | + | + | Low (36.4) |

*Note.* (++) = yes; (+) = partially yes; (–) = no; NA = not applicable

^a^Q5-7 not applicable due to observational nature of included studies

**Figure S1**. Study-location subgroup analysis

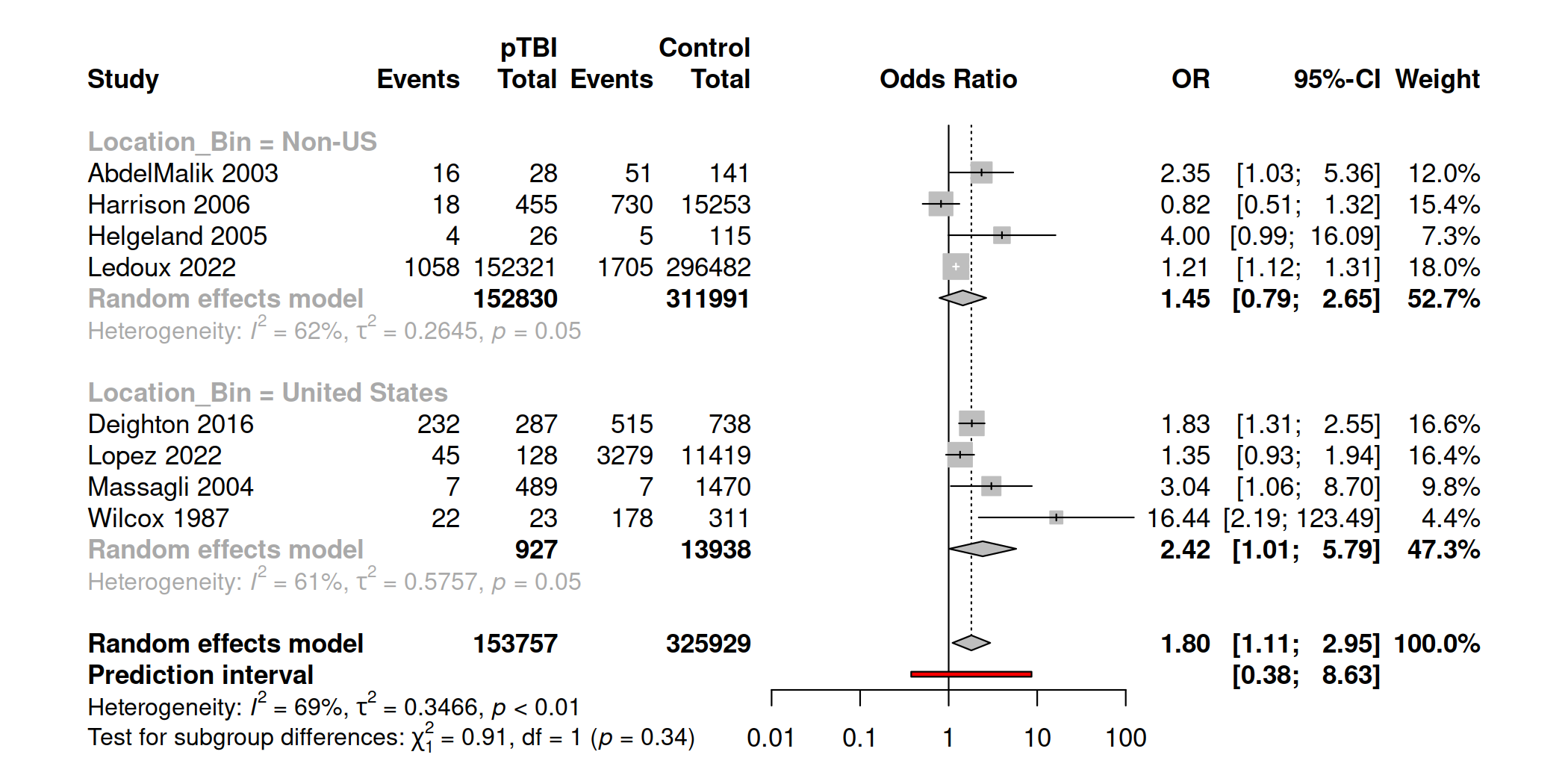

**Figure S2**. Study-design subgroup analysis

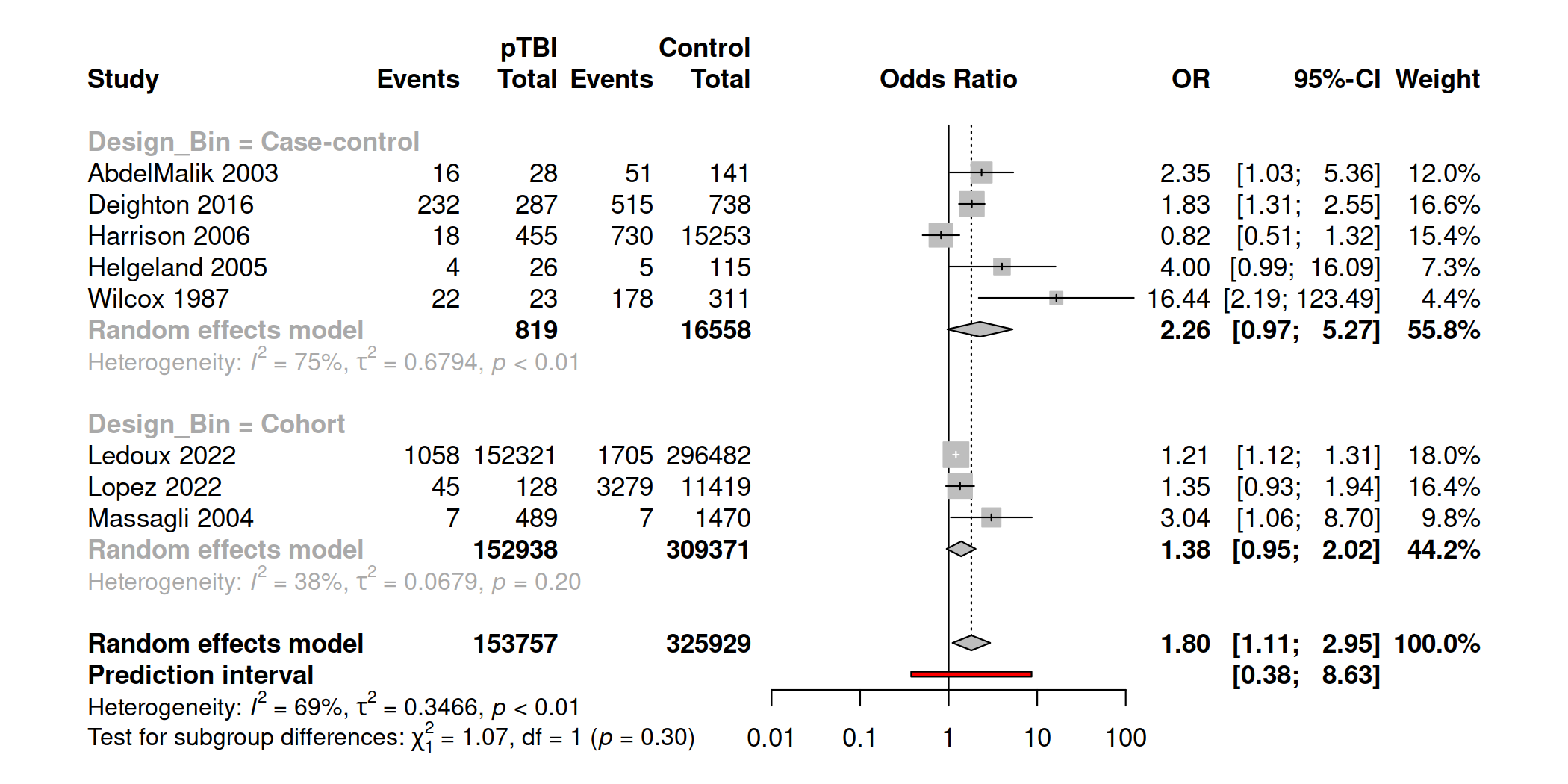

**Figure S3**. Psychotic disorders or sub-threshold symptoms subgroup analysis

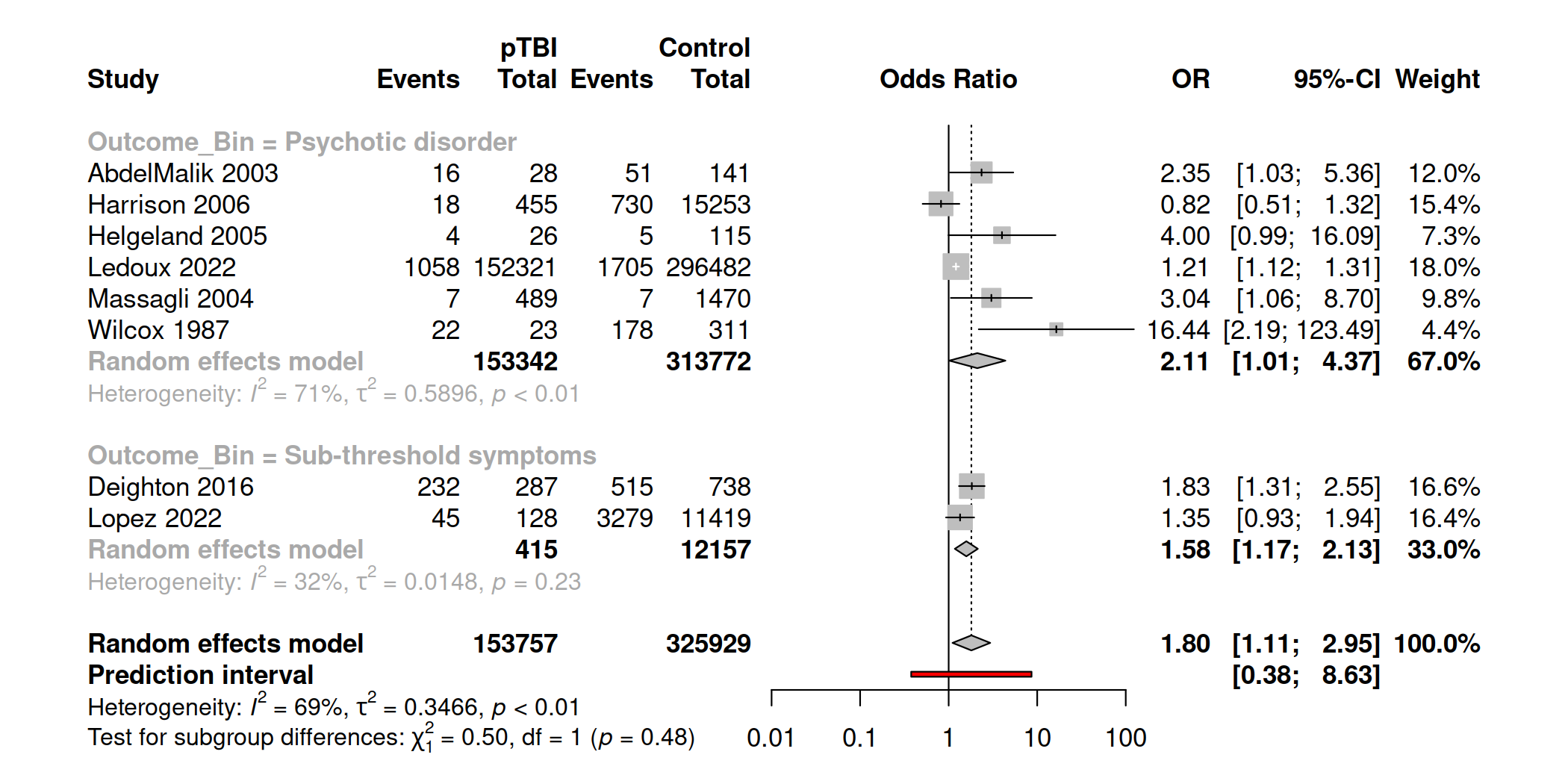

**Figure S4**. Type-of-outcome-measurement subgroup analysis

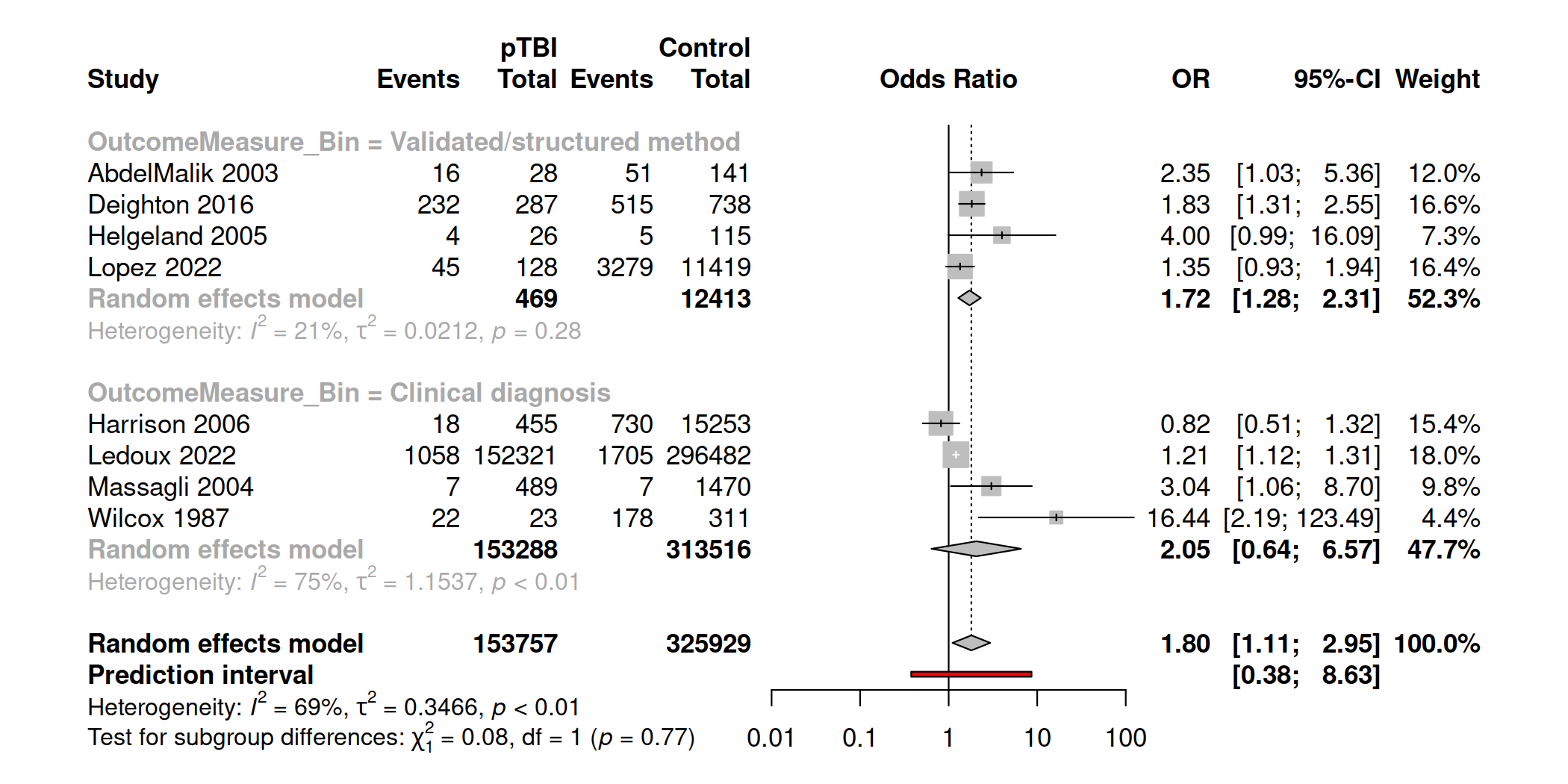

**Figure S5**. Onset-time-of-outcome subgroup analysis

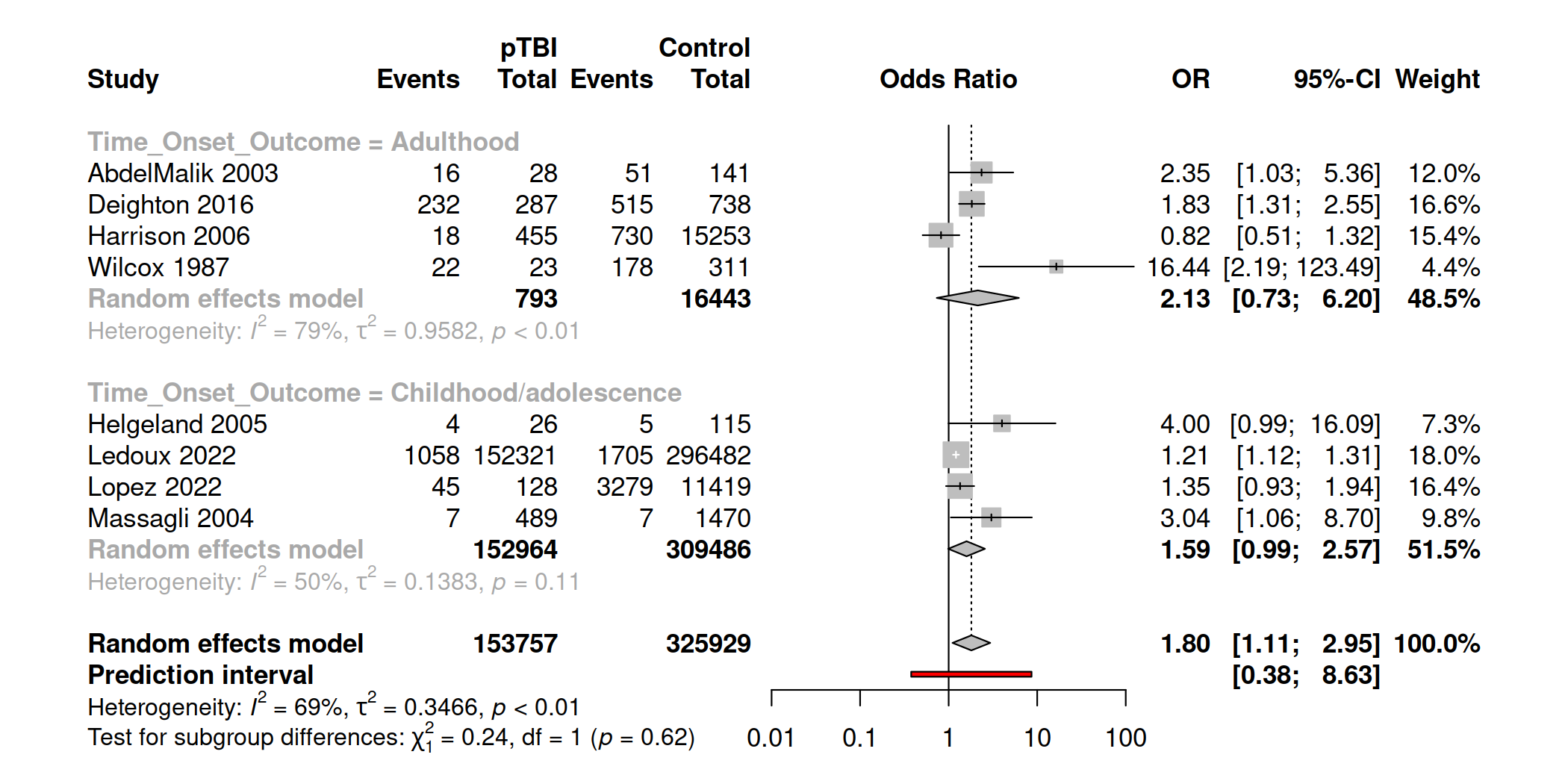
